# Community occurrence of metapneumovirus, influenza A, and respiratory syncytial virus (RSV) inferred from wastewater solids during the winter 2022-2023 tripledemic

**DOI:** 10.1101/2023.06.12.23291120

**Authors:** Alexandria B. Boehm, Marlene K. Wolfe, Bradley White, Bridgette Hughes, Dorothea Duong, Amanda Bidwell

## Abstract

Wastewater monitoring can provide insights into respiratory disease occurrence in communities that contribute to the wastewater system. Using daily measurements of RNA of influenza A (IAV), respiratory syncytial virus (RSV), and human metapneumovirus (HMPV), as well as SARS-CoV-2 in wastewater solids from eight publicly owned treatment works in the Greater San Francisco Bay Area of California between July 2022 until early May 2023, we identify a “tripledemic” when concentrations of IAV, RSV, and SARS-CoV-2 peaked at approximately the same time. HMPV was also widely circulating. We designed novel hydrolysis probe RT-PCR assays for different IAV subtype makers to discern that the dominant circulating IAV subtype was H3N2. We show that wastewater data can be used to identify onset and offset of wastewater disease occurrence events that can provide insight into disease epidemiology and timely, localized information to inform hospital staffing and clinical decision making to respond to circulating viruses. Whereas RSV and IAV wastewater events were mostly regionally coherent, HMPV events displayed localized occurrence patterns.

## Introduction

Acute respiratory illness (ARI) accounts for a large burden of infectious disease^1,2^; and is a leading cause of death for children under 5 globally ^3^. ARI surveillance in the United States (US), and much of the world, is passive, relying on institutions to identify specific diseases through clinical specimens and regularly reporting results to a government agency. As a result, ARI surveillance is biased towards identifying infections in individuals with co-morbidities and/or where symptoms are severe^4^. Resultant lack of knowledge on occurrence and trends in circulating respiratory disease dynamics limits institutional awareness to guide public health response and clinical decision making. Additionally, the lack of robust, unbiased data on disease occurrence limits efforts to understand disease epidemiology.

During the COVID-19 pandemic, the circulation of respiratory viruses changed dramatically according to both clinical surveillance^5,6^ and wastewater monitoring^7^. For example, in the US, there was limited influenza or RSV circulation in the northern hemisphere winter of 2021- 2022^5,8^. During winter 2022-2023, the US experienced a “tripledemic” of respriatory viruses as clinics and hospitals recorded large numbers SARS-CoV-2, influenza, and RSV infections, at times overwhelming hospital capacity particularly in pediatric units^9^. Nearly 40% of United States households reported being infected by SARS-CoV-2, influenza, or RSV during the tripledemic period^10^.

In the present study, we explore the potential for wastewater monitoring, also referred to as wastewater-base epidemiology, to inform the onset and progression of important respiratory viruses during this tripledemic period in the United States. We measured concentrations of genomic nucleic-acids of influenza A (IAV), RSV, SARS-CoV-2, as well as human metapneumovirus (HMPV) in wastewater solids daily in eight publicly owned treatment works (POTWs) in the Greater San Francisco Bay Area of California, USA during winter 2022-2023. We find that the unusual co-occurence of the onset of these outbreaks is identifiable using wastewater monitoring, and moreover HMPV was also circulating. Whereas clinical testing data are affected by an individual’s test seeking behavior and test availability^11–13^, as well as institutional reporting delays which can be weeks in duration^14,15^, wastewater data are available within 24 hours of sample collection and provide information about the entire community contributing to the wastewater system. We posit that wastewater can be used to help identify the onset and offset of respiratory virus transmission events, as well as the peak of such events. This in turn can inform clinical decision making as well as institutional public health messaging and individual behaviors.

## Methods

### Sample collection

Eight POTWs located in the greater San Francisco Bay Area (extending to Sacramento) in California, USA contributed daily samples of wastewater settled solids for the prospective study (Figure S1, additional POTW details are in Wolfe et al.^16^). Approximately 50 ml of settled solids were collected daily between 7/1/22 (month/day/year format) and 5/7/23. Additional details of sample collection are in the SI. Samples were immediately stored at 4°C and transported to the laboratory where processing began within 6 h of collection. 2472 samples were collected.

### Sample preparation and RNA extraction

Details of sample preparation and nucleic-acid extraction have been described in detail elsewhere^7,16–18^ and are described briefly in the SI. RNA was extracted from 10 replicate aliquots per sample and then subjected to an inhibitor removal step. Extraction-negative controls (water) and extraction-positive controls were extracted using the same protocol as the homogenized samples. The positive controls consisted of SARS-CoV-2 genomic RNA (ATCC VR-1986D), Twist Synthetic Influenza A H3N2 RNA Control (Twist 103002), Intact RSV B virus (Zepto NATFVP-NNS), and a gene block (double-stranded DNA [dsDNA] purchased from IDT) for the HMPV target in the BCoV-spiked DNA/RNA shield solution described above.

### Droplet digital PCR

RNA extracts were used as template in digital droplet RT-PCR assays for PMMoV, BCoV, SARS-CoV-2 N, IAV M, RSV N, and HMPV L gene targets in multiplex assays which have been previously published^7,19^. Each of the 10 replicate nucleic-acid extracts were run in their own well. Assays were multiplexed using the probe mixing approach (see SI and Table S1). PMMoV is highly abundant in wastewater globally^20^ and is used here as an internal recovery and fecal strength control^21^. Undiluted extract was used for the human viral assay template, and a 1:100 dilution of the extract was used for the BCoV/PMMoV assay template. Details of the digital RT-PCR have been published and are included in the SI.

Concentrations of RNA targets were converted to concentrations per dry weight of solids in units of cp/g using dimensional analysis. The total error is reported as standard deviations and includes the errors associated with the Poisson distribution and the variability among the 10 replicates. BCoV recovery was determined by normalizing the BCoV concentration by the expected concentration given the value measured in the spiked DNA/RNA shield. BCoV recovery was used as a process control and not used in the calculation of concentrations; samples were rerun in cases where the recovery of BCoV was less than 1%.

### Influenza subtypes

We designed and tested novel RT-PCR primers and internal hydrolysis probes targeting the H1 subtype of the hemagluttanin (HA) gene, H3 subtype of the HA gene, N1 subtype of the neuraminidase (NA) gene, and N2 subtype of the NA gene of IAV. Primers and probes were then screened for specificity in silico, and in vitro against other respiratory and enteric viruses (see SI, Table S2).

We retrospectively tested two samples per week between 7/1/22 and 3/31/23 (covering most of the IAV wastewater event) from SJ POTW for the four influenza A subtype markers (N1, N2, H1, H3). We also re-analyzed the samples to quantify the IAV M gene in order to report the ratio of the subtype markers to M gene (see SI).

### Clinical data

We used state-aggregated weekly clinical sample positivity rates from sentinel laboratories for IAV, RSV and HMPV. IAV, RSV, and a fraction of the HMPV data are publicly available^22^, the remaining HMPV data were provided by California Department of Public Health (CDPH). State-aggregated daily positivity rates for COVID-19 are publicly available (https://data.chhs.ca.gov/dataset/covid-19-time-series-metrics-by-county-and-state).

### Data analysis

Human viral RNA concentrations were normalized by PMMoV RNA; 5-d trimmed averages were used to visualize data and identify the date of their peak maximum concentrations. The onset dates of IAV, RSV, and HMPV wastewater events were identified as the first day for which all samples in a 14-d look back period had concentration higher than 2000 copies/g,, which is approximately twice the lowest detectable concentration. The offset dates of IAV, RSV, and HMPV wastewater events were identified as the first day after an onset event for which only 7 samples during a 14-d look back period had concentrations over 2000 copies/g.

We tested whether PMMoV-normalized viral RNA concentrations at each POTW were correlated to the same measurements at the other eight POTWs (28*4 = 112 hypothesis tests), and whether PMMoV-normalized viral RNA concentrations were correlated within POTWs (48 tests) using Kendall’s tau as these data tend to not be normally distributed. We also tested the hypothesis that PMMoV-normalized viral RNA correlated to state-aggregated positivity rates (32 tests). Since IAV, RSV, and HMPV positivity rates are aggregated weekly, we used weekly median PMMoV-normalized viral RNA concentrations to test the hypothesis. We used a p value of 0.00026 (0.05/192) which corresponds to alpha= 0.05 with a Bonferroni correction. Results were similar when analyses were carried out with variables that were not PMMoV-normalized.

This study was reviewed by the State of California Health and Human Services Agency Committee for the Protection of Human Subjects and determined to be Exempt from oversight. Wastewater data are publicly available (https://purl.stanford.edu/gp563rt4747). Data collected between 7/1/22 and 12/31/22 are publicly available through a Data Descriptor^23^.

## Results

### QA/QC

Results are reported as suggested in the Environmental Microbiology Minimal Information guidelines^24^ (Figure S2). Extraction and PCR negative and positive controls performed as expected (negative and positive, respectively). Median BCoV recovery in these solids samples was 1.2 (interquartile range = 0.92-1.72, n=2472); values greater than 1 are likely a result of uncertainty in quantifying the amount of BCoV spiked in the DNA/RNA shield.

### Novel IAV subtype assay sensitivity and specificity

In silico analysis indicated no cross reactivity of the novel RT-PCR probe-based assays (Table S3) with sequences deposited in NCBI. The novel assays for IAV N1, N2, H1 and H3 were tested in vitro against non-target viral gRNA as well as target gRNA. No cross reactivity was observed.

### Viral RNA in wastewater solids

Viral concentrations were dynamic over the project duration, with most showing elevated concentrations in the winter (Figure 1). PMMoV-normalized HMPV, IAV, RSV, and SARS-CoV-2 RNA concentrations were each positively and significantly associated between the eight POTWs, respectively (IAV tau: 0.49 to 0.64, RSV tau: 0.53 to 0.71, HMPV tau: 0.37 to 0.56, SARS-CoV-2 tau: 0.27 to 0.45, all p<10^-12^, Figure S3) suggesting coarse temporal coherence across the region.

**Figure 1.**
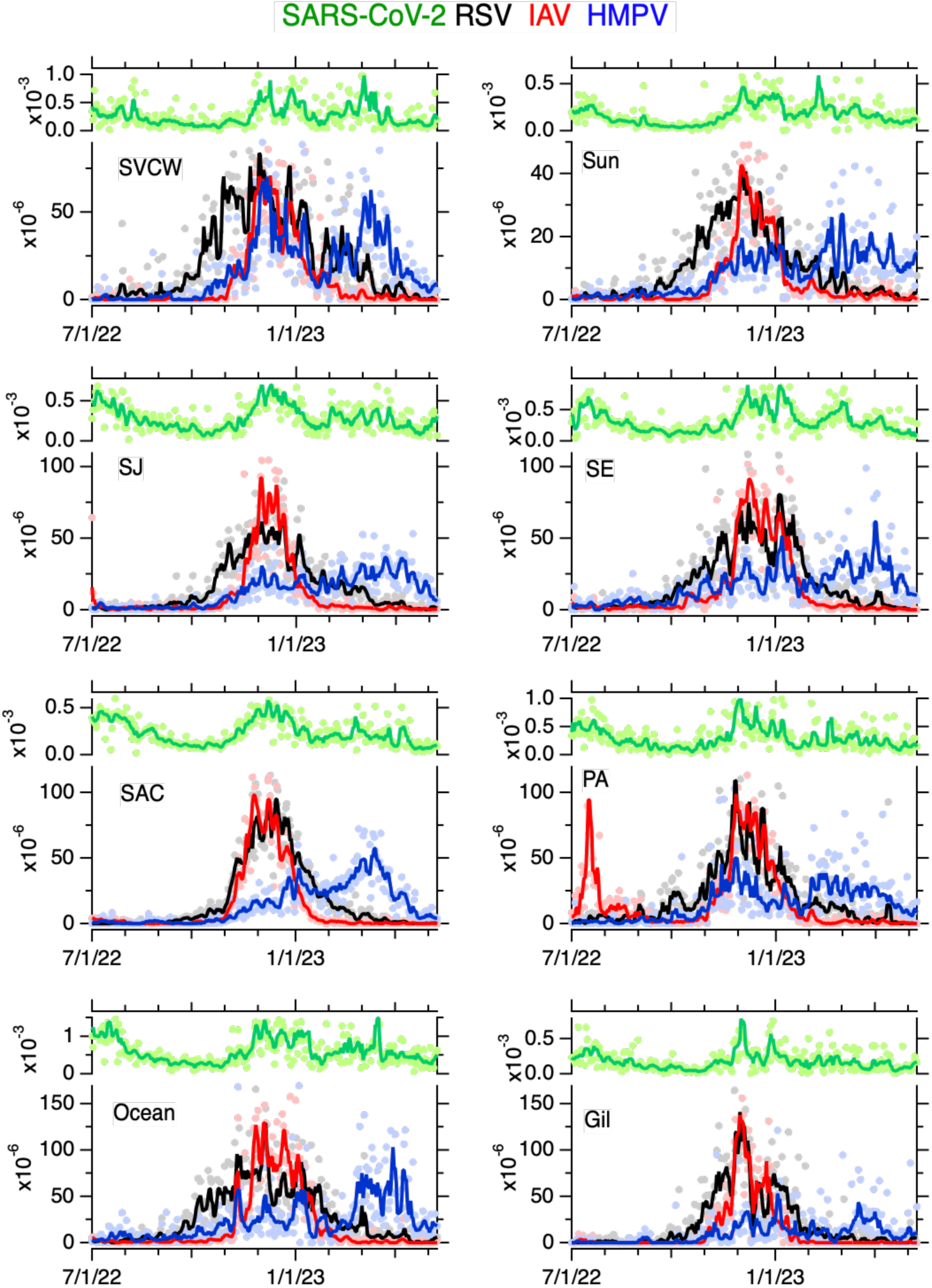
Time series of concentrations of SARS-CoV-2, RSV, IAV, and HMPV normalized by PMMoV in wastewater solids at eight POTWs between 7/1/22 and 5/7/23; the ratio is unitless. Raw data are shown as filled circles, lines are 5-d trimmed averages. Note the difference in scale between the SARS-CoV-2 (upper left axis of each panel) and the other viruses (lower left axis of each panel). The acronym for each POTW is provided in the upper left corner.

Within each POTW, PMMoV-normalized IAV, RSV, HMPV, and SARS-CoV-2 RNA concentrations were generally positively associated with each other suggesting coherence within a POTW (tau: 0.15 to 0.56, all p < 0.00026, depending on POTW and specific measurements, Figure S4). The exceptions are at PA where HMPV was not associated with IAV, at Gil where HMPV was not associated with SARS-CoV-2, and at SAC and Ocean, where HMPV was not significantly associated with IAV or SARS-CoV-2. Taus were generally highest between IAV and RSV, and slowest between HMPV and both IAV and SARS-CoV-2.

State-aggregregated clinical specimen positivity rates for IAV, RSV, HMPV, and SARS-CoV-2 show peaks in winter months (Figure S5). Weekly median PMMoV-normalized concentrations of all viruses were significantly, positively correlated with the associated clinical positivity rate (see SI, all p<10^-6^).

The peak wastewater concentrations varied across POTWs for the different viral targets (Table 1). Peak RSV occurred between 11/25/22 and 12/14/22, peak IAV occurred between 26 Nov and 13 Dec, and peak HMPV occurred between 11/26/22 and 3/30/23, depending on POTW. For context, peak SARS-CoV-2 occurred between 11/30/22 and 3/16/23.

**Table 1.** Date of onset and offset, as well as peak of wastewater events for each human viral target measured in wastewater solids for each of 8 POTWs. A “?” indicates that the offset had not yet occurred during the study period. Dates for two events are provided if two wastewater events were identified. SC2 is SARS-CoV-2, only the peak is provided for SC2.

| POTW | HMPV |  | IAV |  | RSV |  | SC2 |
| --- | --- | --- | --- | --- | --- | --- | --- |
|  | onset/offset | peak | onset/offset | peak | onset/offset | peak | peak |
| Gil | 3 Dec - ? | 3 Jan | 13 Nov - 25 Jan | 30 Nov | 24 Oct - 20 Mar | 30 Nov | 1 Dec |
| SJ | 30 Oct - ? | 22 March | 14 July - 23 July<br>8 Nov - 3 Mar | 30 Nov | 17 Sept - 18 April | 20 Dec | 1 Dec |
| PA | 1 Nov - ? | 26 Nov | 14 July - 21 Sept<br>11 Nov - 13 Feb | 26 Nov | 11 Oct - 2 April | 25 Nov | 30 Nov |
| SVCW | 12 Nov - ? | 6 Dec | 4 Nov - 24 Feb | 9 Dec | 23 Sept - 25 April | 13 Dec | 4 Mar |
| SE | 1 Sept - 24 Sept<br>13 Oct - ? | 1 April | 26 Oct - 25 Feb | 8 Dec | 6 Oct - 25 Mar | 29 Nov | 5 Jan |
| Ocean | 6 Aug - ? | 30 March | 18 Nov - 22 Feb | 13 Dec | 23 Sept - 17 April | 4 Dec | 16 Mar |
| SAC | 10 Nov - ? | 13 March | 4 Nov - 30 Jan | 24 Nov | 19 Oct - 27 Mar | 14 Dec | 7 Dec |
| Sun | 4 Nov - ? | 2 March | 10 Nov - 1 Mar | 2 Dec | 26 Sept - 29 April | 3 Dec | 9 Feb |

We identified dates of wastewater event onsets and offsets for RSV, IAV, and HMPV, but not for SARS-CoV-2 as its levels were such that the entire period of the study would be defined as a wastewater event (Table 1). We observed wintertime RSV wastewater events at all 8 POTWs with onsets between 9/17/22 and 10/24/22, and offsets between 3/20/23 and 4/29/23, depending on POTW. The period of time between onset and offset varied between 147 and 215 days (median = 190 days).

We observed a summer IAV wastewater event at two POTWs (SJ and PA); both onset on 7/14/22 and offset on 7/23/22 (SJ) and 9/21/22 (PA). We observed wintertime wastewater events at all 8 POTWs with onsets between 10/26/22 and 11/18/22 and offsets between 1/25/23 and 3/3/23. The duration of the wintertime events ranged from 73 and 122 days (median = 104 days). IAV subtype analysis at SJ did not detect N1, but detected N2, H1, and H3. When detected, the ratio of N2/IAV ranged from 0.3 to over 1 (median = 0.6), H1/IAV ranged from from 0.03 to 0.35 (median = 0.07), and H3/IAV ranged from from 0.1 to 0.4 (median = 0.2) (Figure S6).

We observed a fall HMPV wastwater event at one POTW (SE) with onset on 9/1/22 and offset on 9/24/22. We observed wintertime HMPV wastewater events for all 8 POTWs with onsets between 8/6/22 and 12/3/22. Data from the 8 POTWs indicated on 5/7/23, the end of the study, that none of the HMPV events had offset suggesting minimal durations (assuming offset occurred on 5/8, the day after the study ended) between 156 and 275 days (median = 187 days).

## Discussion

Influenza A, RSV, and SARS-CoV-2 RNA in wastewater solids in the Greater San Francisco Bay Area of California are reflective of the winter 2022-2023 “tripledemic” when cases of influenza, RSV, and COVID-19 increased dramatically with similar onset periods, sometimes overwhelming hospital capacity^9^. Wastewater monitoring of human metapneumovirus (HMPV)

RNA suggests HMPV was also circulating in communities during the tripledemic, but with slightly different outbreak timing and continued activity after IAV and RSV offset in most areas. HMPV was discovered in 2001^25^ and clinical presentation can be indistinguishable from influenza and RSV^26^. Hospitalization rates associated with HMPV for children are similar to those for influenza^27^. Although there are vaccines available for IAV^28^ and the first vaccine was approved by the FDA for RSV in 2023 (although only for adults age > 60 yo)^29,30^, there is no HMPV vaccine currently available^31^. IAV subtype analysis suggested that H3N2 was more common than H1N1, consistent with subtyping of clinical samples^22^.

Wastewater data was able to resolve information on occurrence patterns at higher resolution than clinical data, showing differences in occurrence patterns of IAV, RSV, SARS-CoV-2, and HMPV community infections. For IAV, we identified two localized summertime events at two POTWs located 15 km apart in Santa Clara County and a late localized fall HMPV event was identified at one POTW located in San Francisco County, neither of which were reflected in their respective state-aggregated positivity rate data.

Wintertime wastewater events were evident for IAV, RSV, and HMPV, but their characteristics differed. IAV events were temporally coherent throughout the region with onsets, peaks, and offsets occurring at all POTWs within the same three, three, and five week windows, respectively. RSV events were also regionally temporally coherent; all POTWs had onsets, peaks, and offsets within the same five, three, and five week windows, respectively. However, wintertime RSV onset and offsets occurred one month earlier and later, respectively, than those of IAV across all POTWs, a pattern also discernible in the state-aggregated clinical data. RSV peak concentrations occurred at the same time as peak IAV concentrations. SARS-CoV-2 concentrations also had local peaks at the same time as RSV and IAV suggesting community infections reach their height in unison, consistent with reports of the tripledemic overwhelming some hospital capacities^9^.

HMPV wastewater events differed widely in the timing of onset and peak among the eight POTWs, suggesting localized dynamics; onsets dates varied by up to 4 months and the timing of the peak varied by up to 5 months. Localized HMPV dynamics is further supported by the relatively weak correlations between HMPV concentrations and concentrations of the other viruses that showed patterns of regional coherence. Although HMPV concentrations were positively associated with the state-aggregated positivity rate data, there are clearly localized dynamics in the wastewater events that are not reflected in the state-aggregated clinical data. More localized data on HMPV circulation is not available as testing is extremely limited^7^. Previous studies of clinical HMPV infections suggest variable seasonal infection from year-to-year in contrast to typical patterns in seasonal RSV and IAV infections^27^.

A global systematic review^32^ indicates that seasonal IAV, RSV, and HMPV epidemics typically overlap. RSV epidemics typically start earlier than IAV epidemics by 0.3 months in temperate regions^32^. In our study RSV wastewater events onset 1.2 months (median) before IAV. The same review indicates IAV and RSV epidemics in temperate areas are 3.8 and 4.6 months in duration, respectively. Wastewater events for IAV and RSV were 3.5 and 4.8 months in duration (medians). Temperate HMPV epidemics began 1.7 months after RSV and were 4.8 months in duration. In the present study, HMPV wastewater events began 1 month after RSV (median), and were at least 6.3 months in duration.

While the clinical test positivity data used here provides insight into disease circulation during the study time period, there is no data on community incidence or prevalence of IAV, RSV, or HMPV in the study area. Additionally, data on COVID-19 incidence and prevalence degraded during this time period owing to the wide availability of at home rapid tests^12,33^, the results of which are not reportable to public health agencies in our study area. IAV, RSV, HMPV, and COVID-19 test positivity rates serve as proxies for trends in disease occurrence, but it is important to acknowledge that the rates reflect those of severely symptomatic cases. There is also a significant delay in those data being available, usually with results available and updated 2-4 weeks after specimen collection. Wastewater represents composite biological samples from an entire community including those who are asymptomatic or mildly symptomatic. Wastewater therefore compliments data on clinical test positivity. Given the delays associated with the positivity rate data, and the sparseness of localized positivity rate data, wastewater can serve as an indicator of the first onsets of localized, community infections. While spatial and temporal coherence was observed for IAV and RSV events, HMPV showed more localized dynamics.

This has been shown previously for other diseases; localized dynamics of mpox were also evident from wastewater monitoring^34^. We explored other methods of identifying wastewater events including using different concentration thresholds and look-back periods. All gave similar relative results among the viruses; a public health organization could decide on data analysis methods for identifying wastewater events that were fit for their purposes.

## Data Availability

Wastewater data are publicly available (https://purl.stanford.edu/gp563rt4747).

## Supporting Information

Additional methodological details as well as Tables S1-S3, and Figures S1-S6.

## Acknowledgements

We acknowledge the numerous people who contributed to wastewater sample collection and Allegra Koch for her support with the literature review. This research was performed on the ancestral and unceded lands of the Muwekma Ohlone people. We pay our respects to them and their Elders, past and present, and are grateful for the opportunity to live and work here.

## Supporting information for

### Additional details of sample collection

At seven of the eight POTWs, settled solids were collected from the primary clarifier. Settled solids samples were grab samples except for at SJ where staff collected a 24-h composite sample^1^. At Gil, solids were settled from a 24-h composite influent sample using standard method 160.5^2^.

### Additional details of sample processing and nucleic-acid extraction

Briefly, solids were dewatered by centrifugation, and an aliquot of dewatered solids was dried to determine its dry weight, and another aliquot was resuspended in the bovine coronavirus (BCoV)-spiked DNA/RNA shield (Zymo Research) at a concentration of 75 mg/ml. Bovine coronavirus (BCoV) was used as a positive recovery control. This concentration of solids was chosen as previous work titrated solutions with various concentrations of solids to identify a concentration that minimized inhibition while maintaining sensitivity of the assays^3,4^. The suspension was homogenized, and then centrifuged. The supernatant was subjected to nucleic acid extraction.

### Additional details of digital droplet RT-PCR

Digital droplet PCR was performed on 20-μl samples from a 22-μl reaction volume, prepared using 5.5-μl template, mixed with 5.5 μl of One-Step RT-ddPCR Advanced kit for Probes (catalog no. 1863021; Bio-Rad), 2.2 μl reverse transcriptase, 1.1 μl dithiothreitol (DTT), and primers and probes at a final concentration of 900 nM and 250 nM, respectively. Droplets were generated using the AutoDG Automated Droplet Generator (Bio-Rad). PCR was performed using Mastercycler Pro with the following protocol: reverse transcription at 50°C for 60 min, enzyme activation at 95°C for 5 min, 40 cycles with 1 cycle consisting of denaturation at 95°C for 30 s and annealing and extension at either 59°C or 61°C (for human viruses) or 56°C (for PMMoV/BCoV duplex assay) for 30 s, enzyme deactivation at 98°C for 10 min, and then an indefinite hold at 4°C. The ramp rate for temperature changes was set at 2°C/s, and the final hold at 4°C was performed for a minimum of 30 min to allow the droplets to stabilize.

All samples were processed for HMPV in multiplex as described in the Data Descriptor by Boehm et al.4. For the remaining human viral assay, samples collected between 7/1/22 and 3/11/23, were processed in multiplex as described in the Data Descriptor by Boehm et al.4 Thereafter, for all samples collected between 3/12/23 and 5/7/23 (month/day/year format), assays for SARS-CoV-2 N gene (FAM), RSV (Cy5), and IAV (Cy5.5) were multiplexed using the probe mixing method; the multiplex assay also contained assays for influenza B and norovirus GII for which results are not presented herein. This assay was run at an annealing temperature of 61°C. BCoV and PMMoV were run in a duplex assay.

Each sample was run in 10 replicate wells. On each 96 well plate, extraction-negative controls were run in 3 wells, and extraction-positive controls in 1 well. PCR-positive controls of SARS-CoV-2, IAV, RSV, HMPV, BCoV, and PMMoV were run in 1 well, and no-template controls (NTC) were run in 3 wells. Positive controls consisted of BCoV and PMMoV gene block controls and the same human virus controls described in the main text as positive extraction controls. Results from replicate wells were merged for analysis.

Droplets were analyzed using QX200 or QX600 Droplet Reader (Bio-Rad). Each sample was run in 10 replicate wells on 96 well plates that also contained positive and negative extraction and PCR controls (see SI). Thresholding was done using QuantaSoft Analysis Pro software (Bio-Rad, version 1.0.596). In order for a sample to be recorded as positive, it had to have at least three positive droplets. Three positive droplets correcponds to a concentration between ∼500 and 1000 copies (cp)/g dry weight; the range in values is a result of the range in the equivalent mass of dry solids added to the wells. Any plates for which negative controls were positive or positive controls were negative were discarded and the samples re-processed and rerun.

### IAV subtype assay design and testing

To design the primers and probes, influenza A genome sequences were downloaded from NCBI in January 2023 and aligned to identify conserved regions in the specified regions of the genome. Then primers and probes targeting those conserved regions were developed in silico using Primer3Plus (https://primer3plus.com/) (Table S2).

Primers and probes were then screened for specificity in silico, and in vitro against other respiratory and enteric viruses. including adenoviruses, coronaviruses, metapneumovirus, parainfluenza, RSV, coxsackievirus, echovirus, parechovirus (NATtrol Respiratory Verification Panel NATRVP2.1-BIO and NATtrol EV Panel NATEVP-C viral panels, Zeptometrix, Buffalo, NY), as well as synthetic genomic RNA from influenza A H1N1 and influenza A H3N2 (Twist Bioscience, South San Francisco, CA).

### Measuring IAV subtype markers in wastewater solids

RNA extracts from the two POTWs (SJ and Ocean) were stored at -80°C for 1-5 months. RNA was subjected to a single freeze-thaw and used as template undiluted in digital droplet PCR using the same methods outlined above. Assays were run in multiplex using the probe mixing approach. H1 and N1 were multiplexed in the HEX and Cy5.5 channels, respectively. H3, N2, and IAV were multiplexed in the HEX, FAM/HEX, and ROX channels. Ten replicates were run for each sample, for each sample with the same number of positive and negative extraction and PCR controls per plate as described for the prospective measurements described in the main text. Positive controls consisted of synthetic genomic RNA from influenza H1N1 and H3N2 (Twist). Results are reported as copies per gram dry weight.

### Correlations between wastewater and clinical positivity rates

IAV in wastewater solids was significantly associated with state-aggregated weekly clinical specimen influenza positivity rates at all POTWs (tau between 0.54 and 0.73, all p<10-6) except for SVCW and Sun where the association was not statistically significant. Weekly median normalized RSV in wastewater solids was significantly associated with state-aggregated weekly clinical specimen RSV positivity rates at all plants (tau between 0.59 and 0.67, all p<10-7). Weekly median normalized HMPV in wastewater solids was significantly associated with state-aggregated weekly clinical specimen HMPV positivity rates at all plants (tau between 0.52 and 0.72, p<10-6). Daily normalized SARS-CoV-2 was positively associated with daily state-aggregated COVID-19 positivity rates (tau between 0.27 and 0.48, all p<10-11), as has been shown extensively in previous work.1,5

---

**Figure S1.**
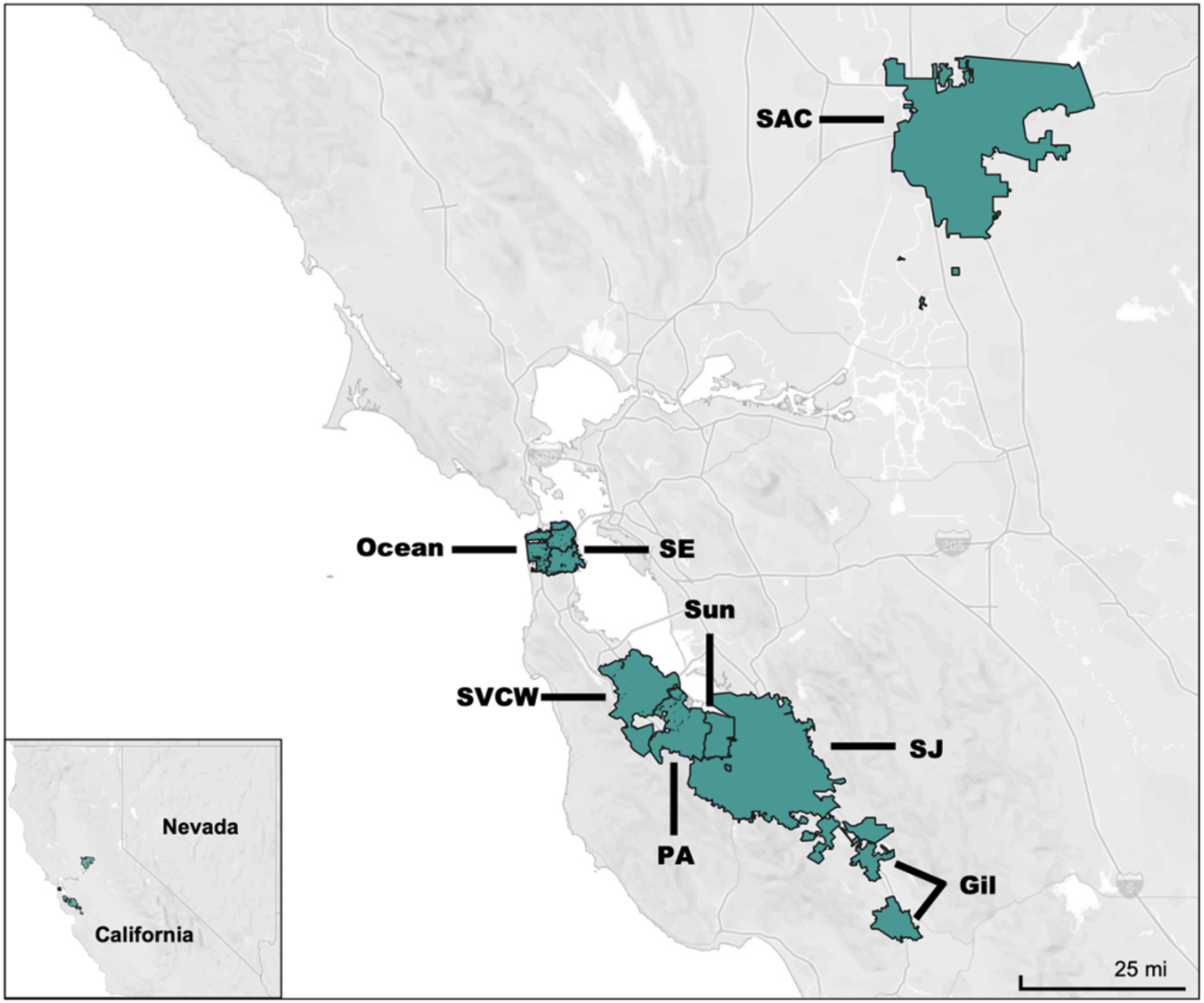
Map of sewersheds participating in the study.

**Figure S2.**
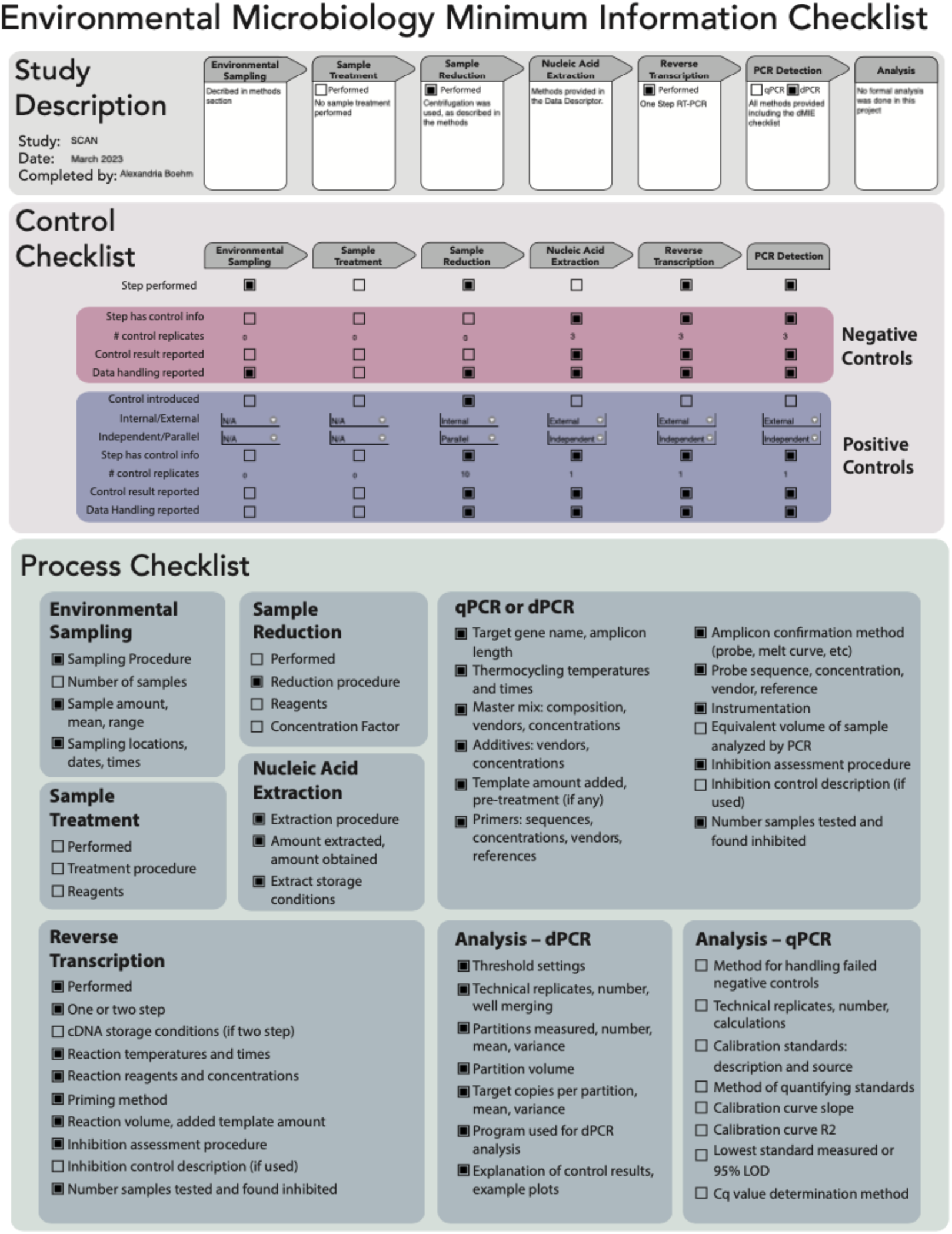
EMMI checklist^6^. Details of the partition numbers and volume, and copy numbers per partition are reported in a Data Descriptor^4^.

**Figure S3.**
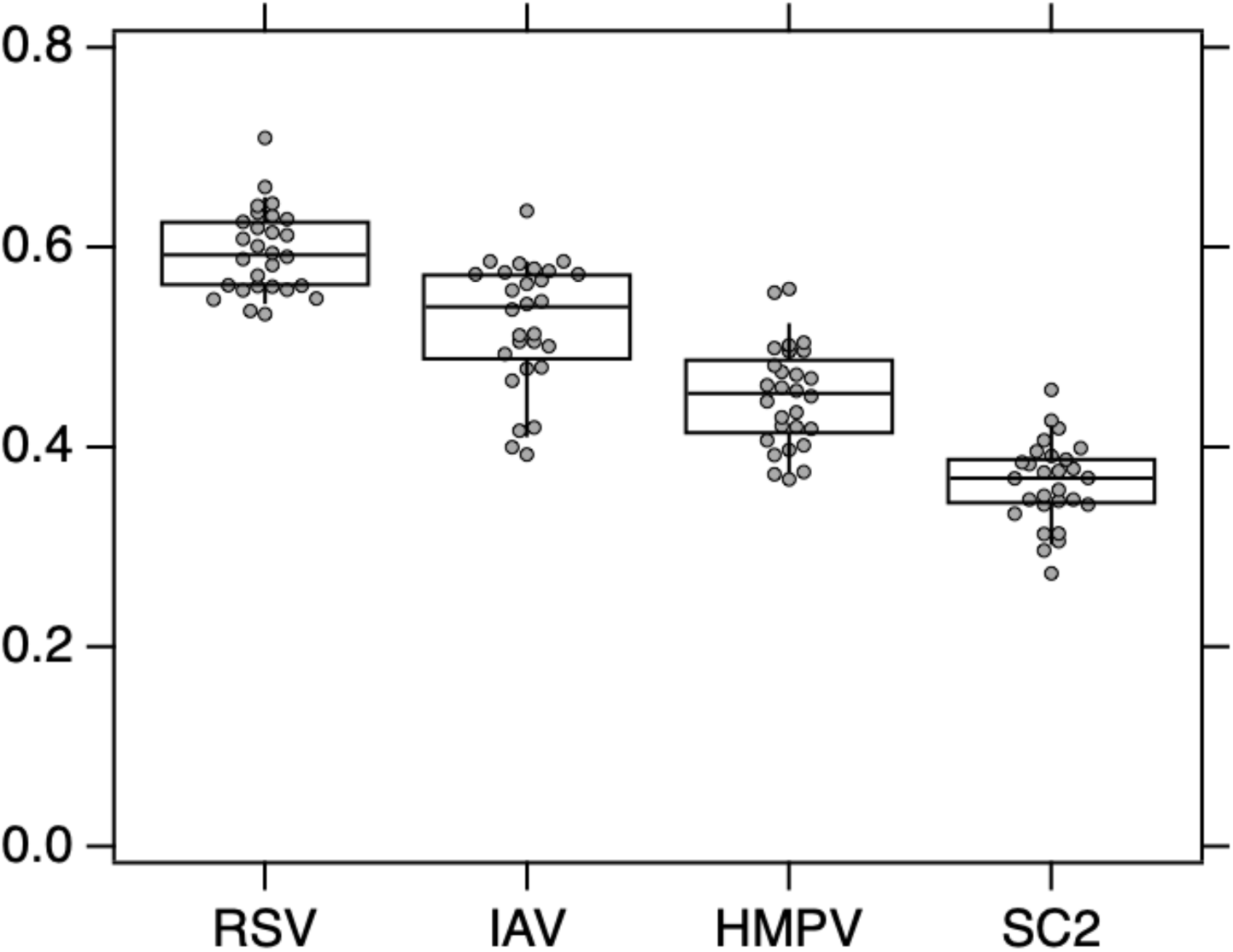
The distribution of between POTW Kendalls tau values for each human viral target. There are 28 tau values for each virus (each combination of 2 POTW among the 8), and those are shown as gray circles. The box and whisker plot provides a visualization of the distribution. The whiskers extend from the 9th to 91st percentiles. The bottom and top of the box represent the 25th and 75th percentiles, the line through the middle of the box represents the median.

**Figure S4.**
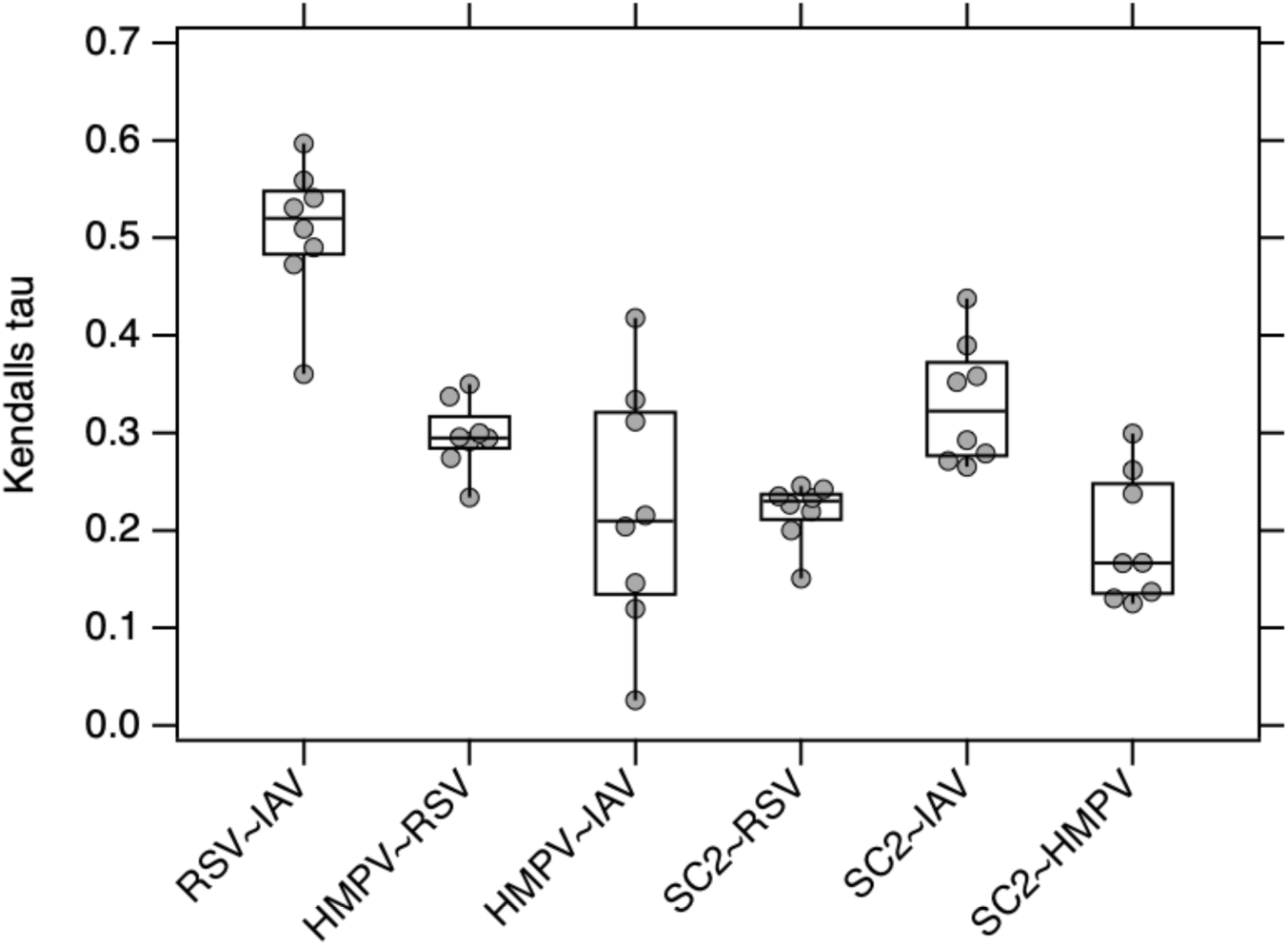
The distribution of within POTW Kendalls tau values for different human viral targets comparisons (bottom axis). There are 8 tau values for each virus combination (1 for each POTW), and those are shown as gray circles. The box and whisker plot provides a visualization of the distribution. The whiskers extend from the 9th to 91st percentiles. The bottom and top of the box represent the 25th and 75th percentiles, the line through the middle of the box represents the median. SC2 is SARS-CoV-2. All tau values are shown, even those that are not significantly different from 0.

**Figure S5.**
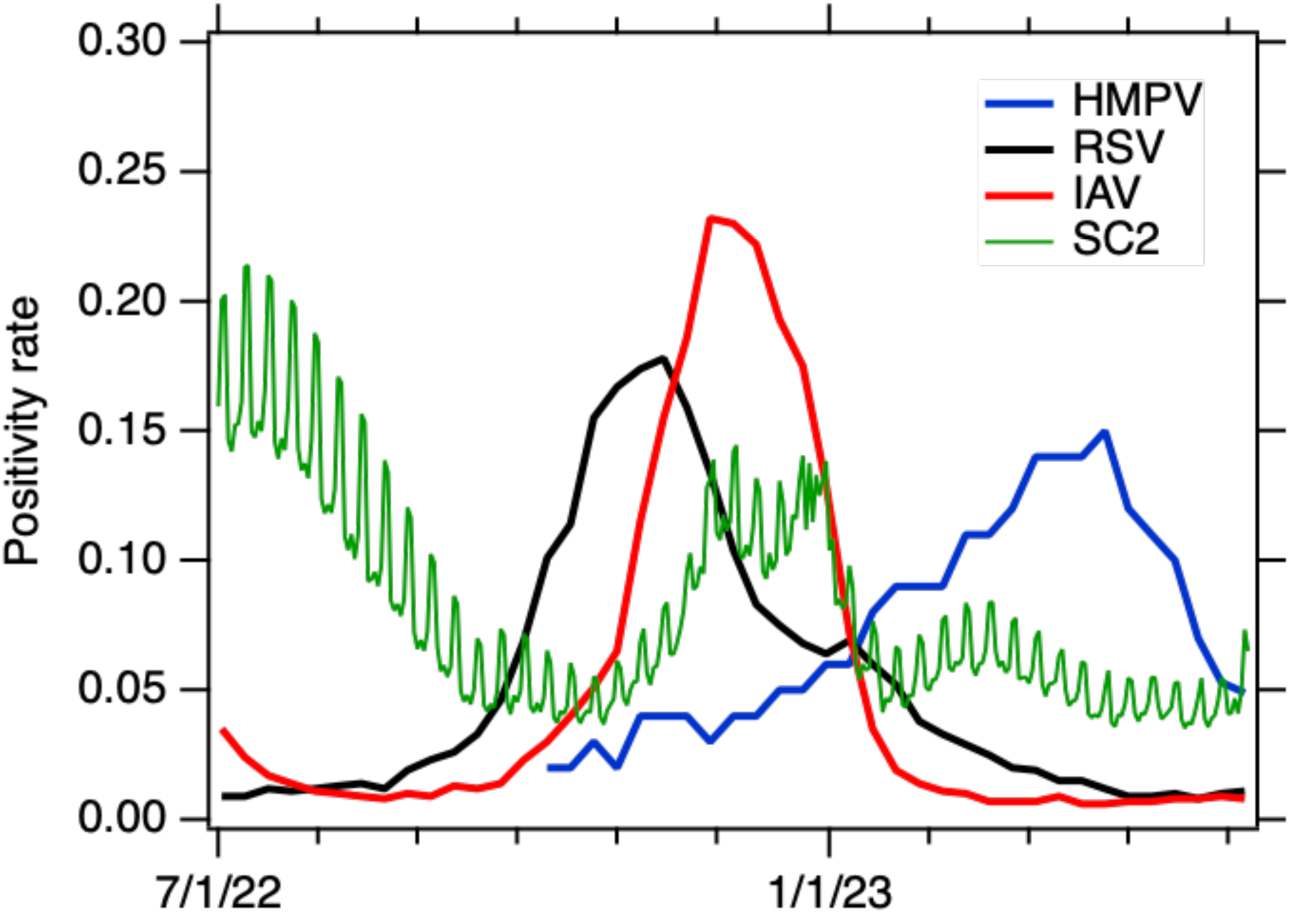
Positivity rates for influenza A (IAV), RSV, human metapneumovirus (HMPV) and SARS-CoV-2 in clinical specimens aggregated across the state of California. Data are reported weekly for IAV, RSV, and HMPV, and daily for SARS-CoV-2.

**Figure S6.**
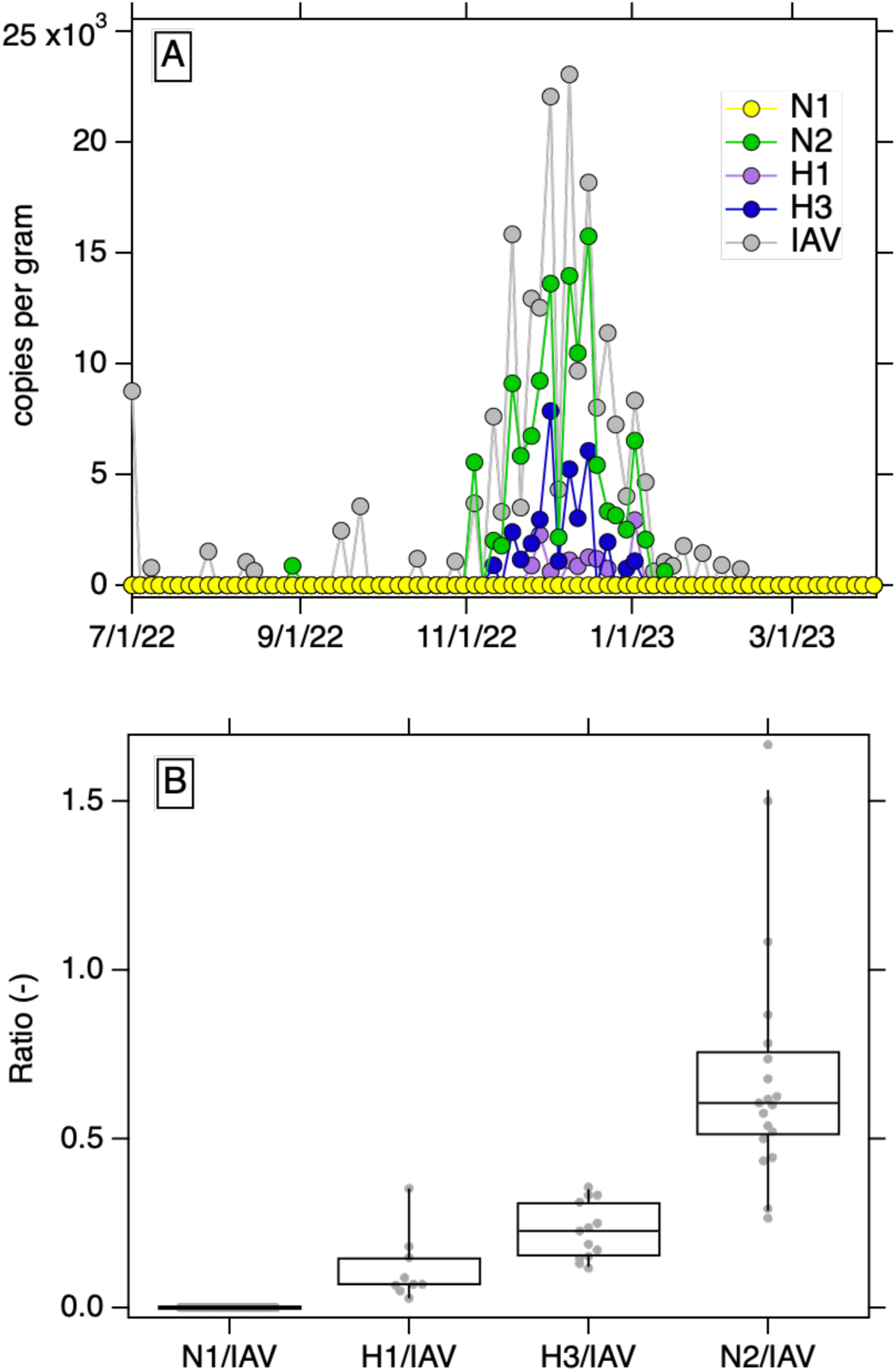
Panel A. Concentrations of IAV, N1, N2, H1, and H3 in copies per gram dry weight in archived samples from SJ POTW. Panel B. Distribitions of the ratio of each IAV subtype marker and IAV measured in the archived samples. Only non-zero ratios are included, aside from N1 for which all values were 0. The whiskers extend from the 9th to 91st percentiles. The bottom and top of the box represent the 25th and 75th percentiles, the line through the middle of the box represents the median.

**Table S1.**
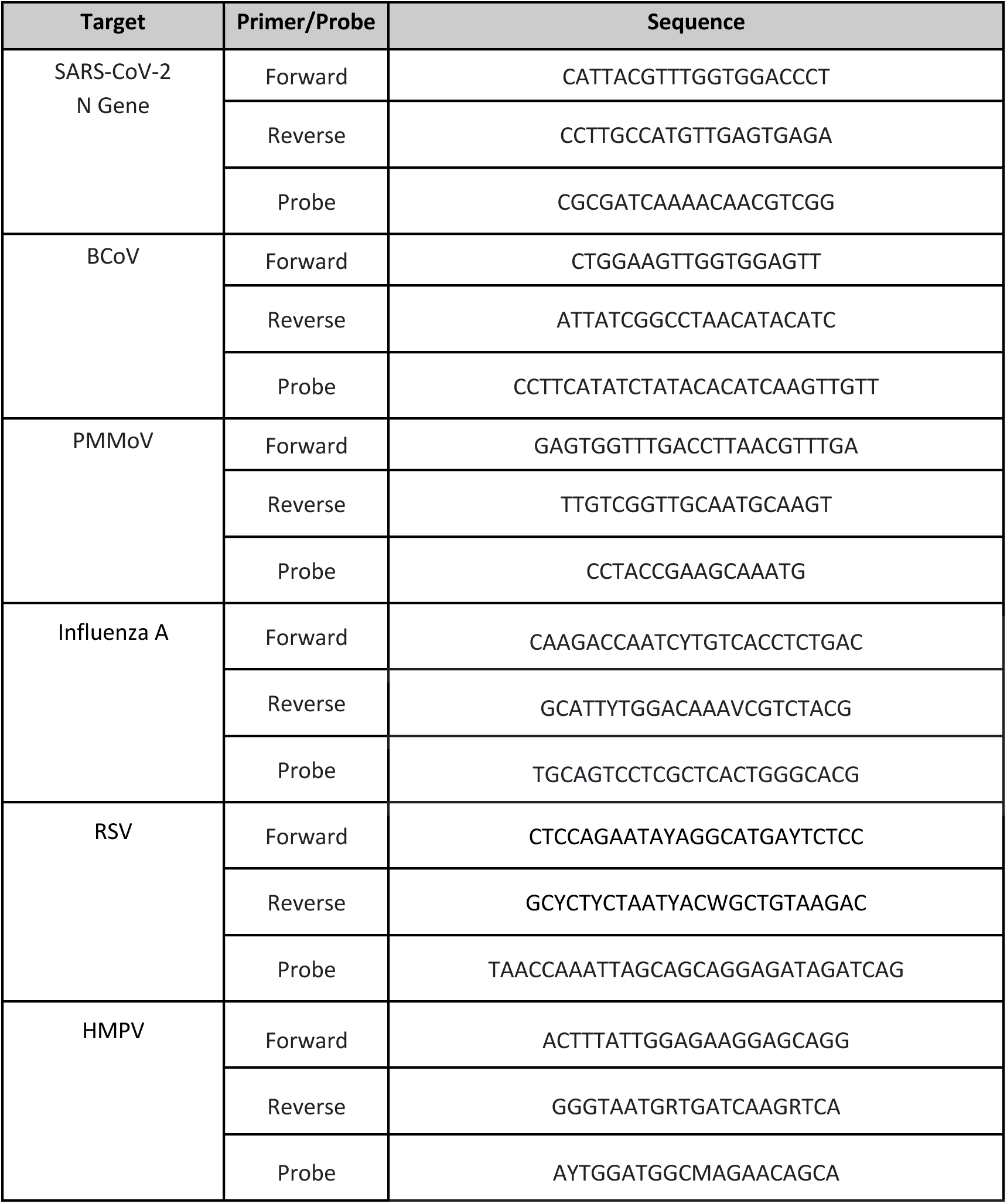
Primers and probes of previously published assays.

**Table S2.**
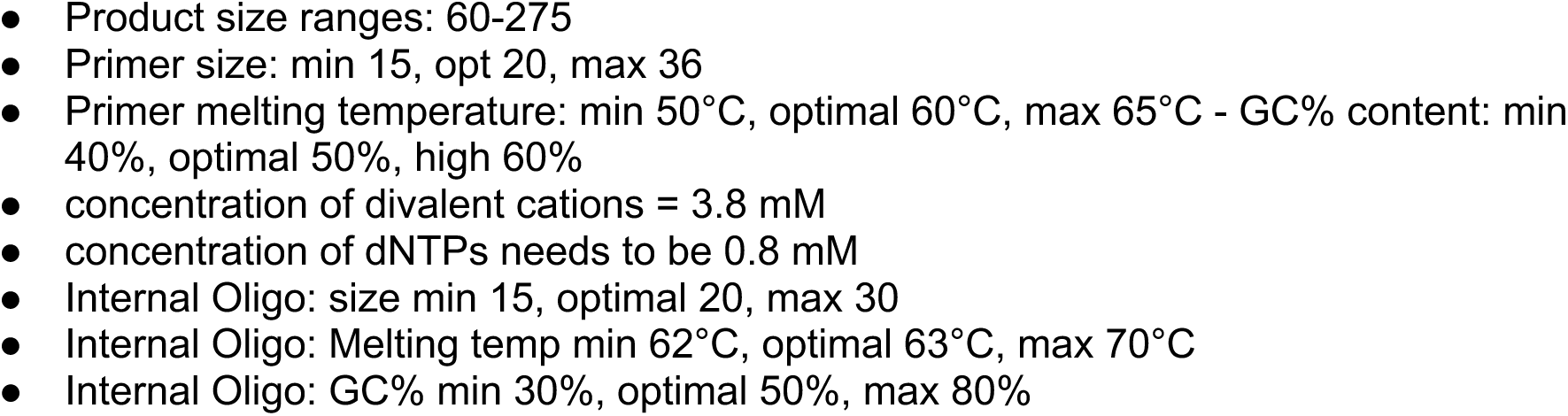
Parameters used with primer design software.

**Table S3.**
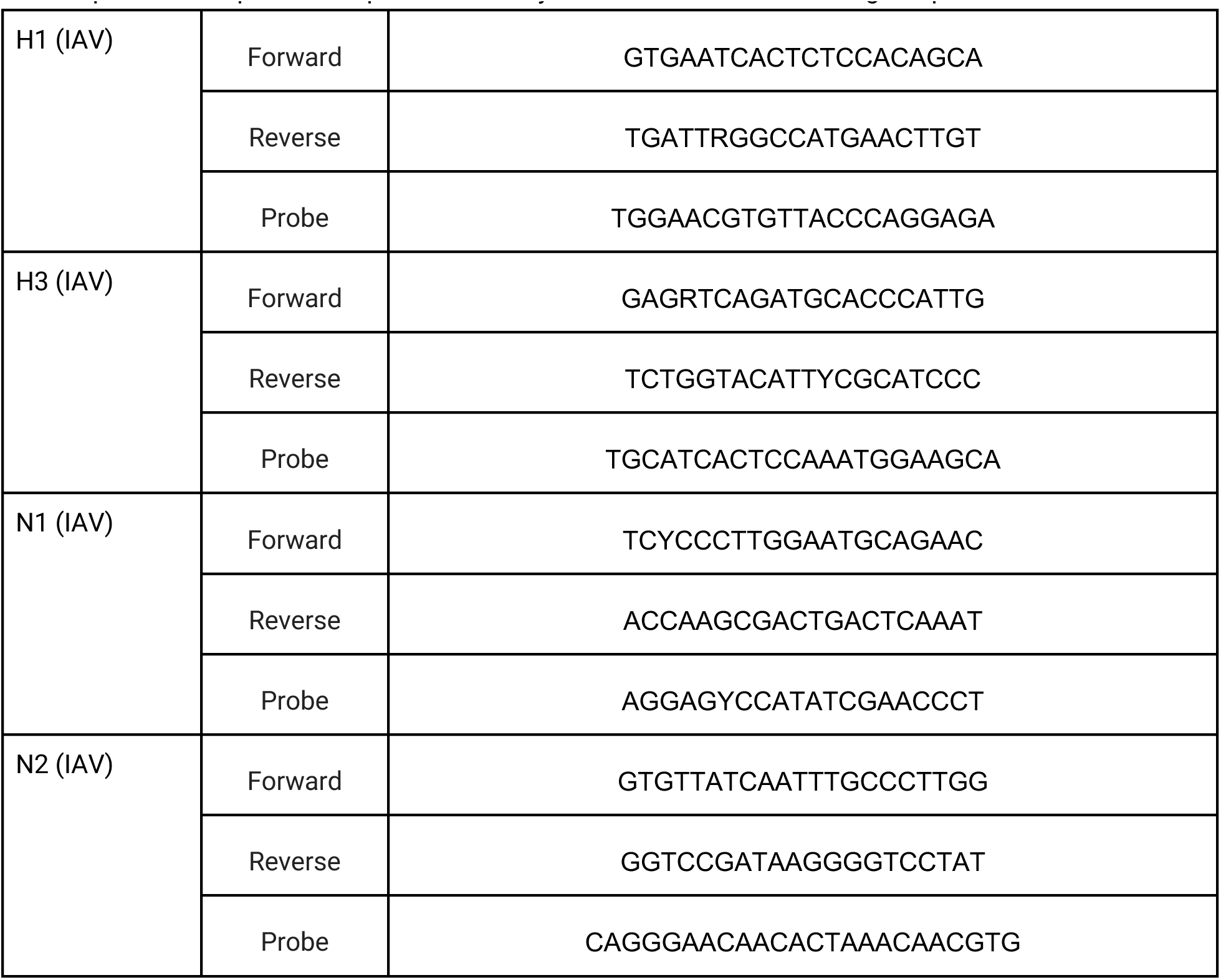
Novel influenza A (IAV) H1, H3, N1, and N2 assays developed in this study. Forward and. reverse primers and probes are provided. Assays were run at 59°C annealing temperatures.

